# Resumption of Sexual Activity After Voluntary Medical Male Circumcision: Data from a Population Based Cohort in Rakai, Uganda, 2013 to 2020

**DOI:** 10.1101/2024.01.03.24300759

**Authors:** Alex Daama, Fred Nalugoda, Edward Kankaka, Asani Kasango, Betty Nantume, Grace Nalwoga Kigozi, Robert Ssekubugu, Juliana Namutundu, Absalom Ssettuba, Tom Lutalo, Joseph Kagaayi, Gertrude Nakigozi, Stella Alamo, Lisa A. Mills, Geoffrey Kabuye, Ron Gray, Maria Wawer, David Serwadda, Nelson Sewankambo, Godfrey Kigozi

**Affiliations:** Rakai Health Sciences Program, Kalisizo, Uganda; Makerere University School of Public Health, Kampala, Uganda; Division of Global HIV & TB, US Centers for Disease Control and Prevention, Kampala Uganda; Johns Hopkins Bloomberg School of Public Health, Baltimore, MD USA

**Keywords:** Trends early sex resumption post-circumcision Uganda, HIV prevention, VMMC, Uganda

## Abstract

**Introduction:** Voluntary medical male circumcision (VMMC) reduces the risk of heterosexual acquisition of HIV by 50%–60%. The Uganda Ministry of Health recommends abstinence for 42 days after VMMC to allow complete wound healing. However, some men resume sex early before the recommended period. We estimated trends in prevalence and risk factors of early sex resumption (ESR) among VMMC clients in Rakai, Uganda, from 2013-2020.

**Methods:** Data from the Rakai Community Cohort Study, a cross-sectional study were analyzed. Data included consenting men aged 15–49 years in who self-reported having received VMMC in one of four successive surveys, (June 2013 to January 2015), (February 2015 to September 2016), (October 2016 to May 2018), and (June 2018 to October 2020). ESR prevalence and associated risk factors using modified Poisson regression to estimate adjusted prevalence ratios (aPR) were estimated.

**Results:** Overall, 1,832 men participated in the study. ESR decreased from 45.1% in 2013 to 14.9% in 2020 (p<0.001). Across the three surveys, ESR prevalence was consistently higher among married than never married men, aPR=1.83, 95% CI: [1.30,2.57]; aPR=2.46, 95% CI: [1.50,4.06]; aPR=2.22, 95% CI: [1.22,4.03]. ESR prevalence was higher among men who reported more than one sexual partner than men with one partner, aPR=1.59, 95% CI: [1.16,2.20]. In the fourth survey (2018-2020), ESR prevalence was significantly higher among men with primary education than men with post-primary, aPR=2.38, 95% CI: [1.31, 4.30] while ESR prevalence was lower among men aged at least 45 men with no sexual relationship in the past 12 months but self-reported to have resumed sex activities, aPR= 0, 95% CI: [1.86-07, 2.69-06] and aPR=0, 95% CI: [3.61e-07, 2.12e-06], respectively. Overall, men who reported primary school as their highest level of education reported ESR more often than those with post-primary education aPR=2.38, 95% CI: [1.31, 4.30]. Occupation and known HIV status were not associated with ESR.

**Conclusions:** Self-reported ESR after VMMC declined between 2013 and 2020. Targeted efforts for counseling focusing on married men, men who had multiple sex partners, and men with lower levels of education can help decrease post-VMMC ESR.

## Introduction

Voluntary medical male circumcision (VMMC) reduces the risk of heterosexual acquisition of HIV by 50%–60%[1][2][3] and was adopted by the Uganda Ministry of Health (MOH) in 2007 following recommendations from the WHO and UNICEF [4] which included guidance that men should abstain from sex for 42 days to allow complete wound healing following VMMC [5][6]

In Uganda, VMMC is provided free of charge with support from the President’s Emergency Plan for AIDS Relief (PEPFAR) through the U.S. Centers for Disease Control and Prevention (CDC). Men undergoing VMMC receive pre-VMMC counseling and follow up 24 hours and 7 days post-VMMC to ascertain wound healing and adherence to 42 days of abstinence. [7] However, previous studies have demonstrated that circumcised men often resume sex early, predisposing them to increased risk of HIV acquisition [8] and increased risk of HIV transmission to their female partners for HIV-positive men[9]. Other risks include wound disruption, bleeding, wound swelling, and pain. Cross-sectional studies indicate that between 25% to 50% of men undergoing VMMC resume sex before the recommended 42 days of abstinence[10][11][12][8]]. Research indicates that factors associated with early sex resumption (ESR) include older age, lower education and unemployment[9], being married[8] and polygamy or multiple sex partners[12]. However, temporal changes in the prevalence of ESR and associated factors are unknown. We estimated prevalence of ESR and associated risk factors among men undergoing VMMC in Rakai, Uganda; over a seven-year period (2013-2020).

## Methods

### Study design and setting

We used cross-sectional data from the Rakai Community Cohort Study (RCCS) and the same standardized questions were asked in all four rounds of the survey. The RCCS is an open population-based cohort which enrolled about 20,000 consenting participants 15-49 years of age in 40 communities across the greater Rakai region. The survey occurs every ∼18 months capturing data on socio-demographic characteristics, sexual behaviors in the past 12 months, HIV, sexually transmitted infections (STIs) including HIV prevention and treatment services, self-reported circumcision status and non-communicable diseases [14]. HIV testing was part of the VMMC program and HIV-positive men were eligible for VMMC. The communities include agrarian, peri urban/trading, and fishing communities along the shores of Lake Victoria. HIV prevalence ranges from 5.2% [15] in agrarian communities to 44.3% in fishing communities [16]. Circumcised men were asked if they resumed sex and if “Yes” when they resumed sex after circumcision (Have you had sex with any one since you were circumcised? If yes, how soon after circumcision did you resume sex?). All variables were self-reported and circumcision status included only men who were medically circumcised. Men who reported being circumcised were asked when they received the service and where they received the service.

### Participants

We included males aged 15-49 years who reported circumcision in any of four successive RCCS survey rounds (June 2013-January 2015), (February 2015-September 2016), October 2016 to May 2018), and (June 2018 to October 2020). Men were censored for all surveys and we considered men who reported recent circumcision (<=3 years). Formal written consent was obtained from each participant prior to data collection. All consenting men who received VMMC services provided blood samples for HIV testing and were archived for future testing. We conducted a multivariate analysis for each survey round.

### Data Analysis

Frequencies and proportions were used to describe participant characteristics by survey round for categorical variables, and means and medians were estimated for continuous variables. At each round, the prevalence and 95% confidence intervals (CIs) were estimated for recently circumcised men reporting early sex resumption < 42 days (ESR). Risk factor analyses was conducted and the dependent variable was ESR; and independent variables included age, number of sexual partners in the past 12 months, education, occupation, marital status, and HIV status. Modified Poisson regression was used to estimate unadjusted and adjusted prevalence ratios (aPR) of ESR at each round. The multivariable model included variables with p-value <= 5% identified during bivariate analyses. Analyses were performed using Stata version 14.0 (StataCorp, 2015). We checked for effect modification including multi-collinearity and found that the mean of VIF of 1.32 hence indicating no relationship between factors.

### Ethical considerations

The RCCS is approved by the Uganda Virus Research Institute (UVRI) Research and Ethics Committee (REC), the Western IRB, and the Uganda National Council Science & Technology. All data were de-identified and participants had provided written informed consent. This project was reviewed in accordance with CDC human research protection procedures and was determined to be research, but CDC investigators did not interact with human subjects or have access to identifiable data or specimens for research purpose.

## Results

### Characteristics of participants by ESR and survey round

Table 1 presents data on participants who were previously circumcised prior to participation in the survey. The prevalence of ESR declined from 45.1% in 2013/2015 to 21.8% in 2015/2016, and 21.8% in 2016/2018 to 14.9% in 2018/2020 [Figure 1]. Prevalence of ESR was higher among men aged 35-44 across three survey rounds; 64.0% in 2013/15, 30.1% in 2015/16 and 25.6% in 2018/20 than other age groups (Table 1).

**Figure I.**
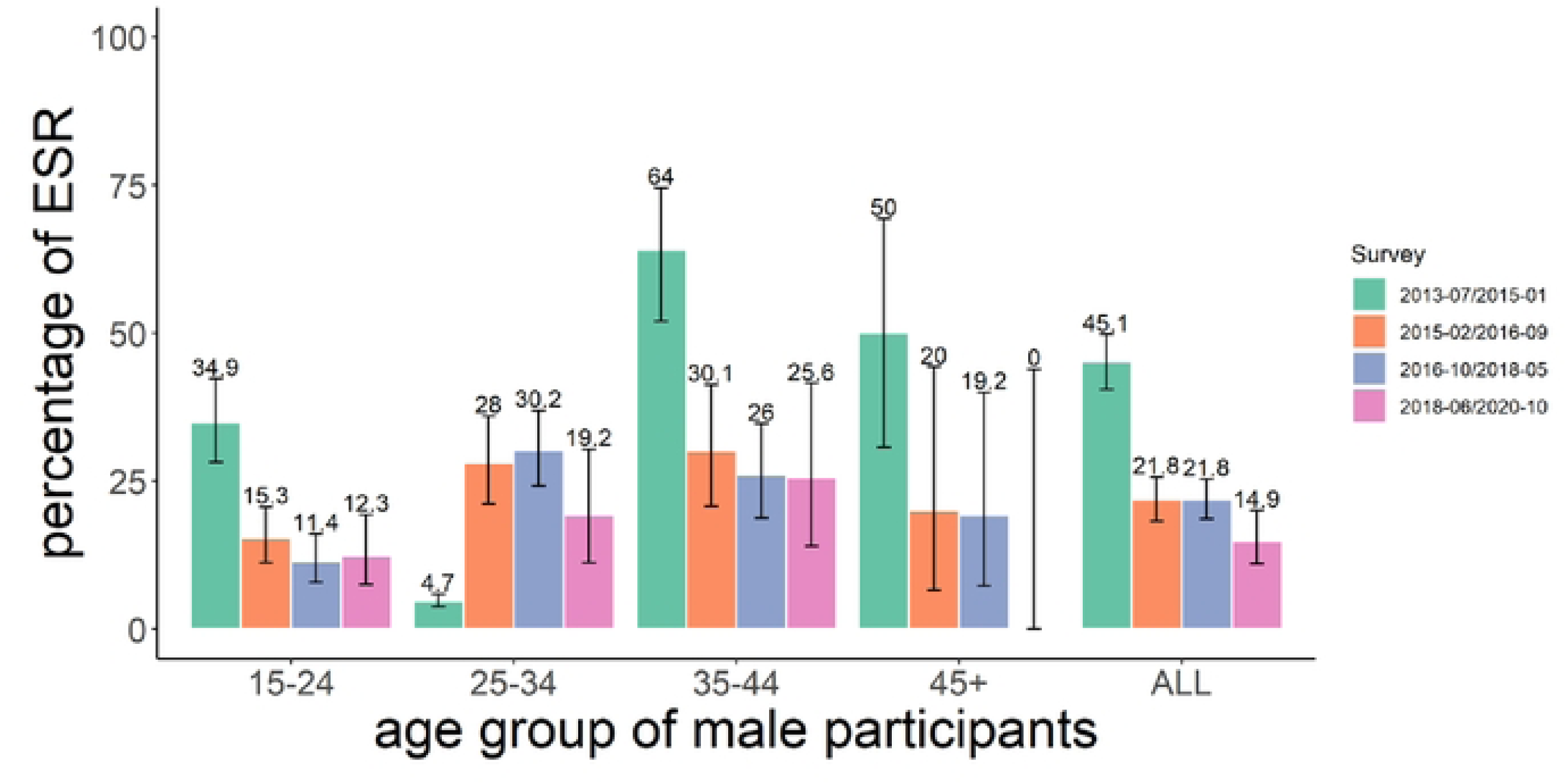
shows early sex resumption following circumcision among men of different age groups over time (2013-2020) in Rakai, Uganda.

**Table 1:** Early sex resumption post circumcision prevalence by survey round and socio-demographic characteristics, 2013-2020, Rakai, Uganda.

| Socio-demographic characteristics | 2013-2015 | 2015-2016 | 2016-2018 | 2018-2020 |
| --- | --- | --- | --- | --- |
|  | This includes the number that resumed sex <42 | This includes the number that resumed sex <42 | This includes the number that resumed sex <42 | This includes the number that resumed sex <42 |

|  | days n=203,<br>Total number<br>resumed N=450<br>n/N (%) | days n=109, Total<br>number resumed<br>N=501 n/N (%) | days n=135,<br>Total number<br>resumed N=620<br>n/N (%) | days n=39 Total<br>number resumed<br>N=261<br>n/N (%) |
| --- | --- | --- | --- | --- |
| Overall prevalence of ESR | 203/450(45.11) | 109/501(21.76) | 135/620(21.77) | 39/261(14.94) |
| Age in years |  |  |  |  |
| 15-19 | 16/60(26.67) | 9/92(9.78) | 7/95(7.37) | 6/71(8.45) |
| 20-24 | 49/126(38.89) | 29/156(18.59) | 22/160(13.75) | 11/67(16.42) |
| 25-34 | 79/167(47.31) | 42/150(28.00) | 64/212(30.19) | 14/73(19.18) |
| 35-44 | 48/75(64.00) | 25/83(30.12) | 33/127(25.98) | 11/43(25.58) |
| 45+ | 11/22(50.00) | 4/20(20.00) | 5/26(19.23) | 0/7(0.00) |
| Marital Status |  |  |  |  |
| Currently married | 135/236(57.20) | 76/233(32.62) | 88/305(28.85) | 22/94(23.40) |
| Previously married | 23/56(41.07) | 7/42(16.67) | 22/77(28.57) | 5/34(14.71) |
| Never married | 45/158(28.48) | 26/226(11.50) | 21/238(8.82) | 15/133(11.28) |
| HIV Status |  |  |  |  |
| Negative | 174/384(45.31) | 89/437(20.37) | 95/509(18.66) | 38/228(16.67) |
| Positive | 29/66(43.94) | 20/64(31.25) | 36/111(32.43) | 4/33(12.12) |
| Number of Sexual Partners (in the past 12 months) |  |  |  |  |
| One partner in the past 12 months | 89/220(40.45) | 51/263(19.39) | 50/313(15.97) | 16/117(13.68) |
| More than one in the past 12 months | 95/187(50.80) | 58/226(25.66) | 80/284(28.17) | 26/130(20.00) |
| Occupation |  |  |  |  |
| Trading | 28/73(38.36) | 18/93(19.35) | 14/82(17.07) | 11/53(20.75) |
| Agrarian | 107/245(43.67) | 59/272(21.69) | 54/319(16.93) | 22/155(14.19) |
| Fishing | 68/132(51.52) | 32/136(23.53) | 63/219(28.77) | 9/53(16.98) |
| Highest Education |  |  |  |  |
| Post primary | 49/129(37.98) | 30/156(19.23) | 27/167(16.17) | 13/133(9.77) |
| Primary | 142/298(47.65) | 78/334(23.35) | 95/429(22.14) | 28/123(22.76) |
| No formal education | 12/23(52.17) | 1/11(9.09) | 9/24(37.50) | 1/5(20.00) |

ESR prevalence was highest among currently married men across the four surveys, compared to men who were divorced or separated (Table 1). ESR was more common among men reporting more than one sex partner than men with one partner. ESR was higher among fisher men compared to all other occupations across the first three survey rounds in 2013/15, 2015/16, and 2016/18. However, in 2018/20 ESR was lower among fishermen than men who reported to be engaged in trading.

ESR prevalence was also higher among those with primary education in two survey rounds, 23.4% in 2015/16 and 22.8% in 2018/20 compared to those who completed post-primary education and men without formal education.

Trends in prevalence of ESR stratified by age across survey periods When stratified by age, similar trends were observed among individuals aged 15-24 years, 35-44 years, and > 45 years where ESR declined over time. However, ESR increased from 4.7% (CI = 3.8-5.9) in (2013/15) to 28.0% (CI=21.1-36.0) in (2015/16) among individuals aged 25-34 years, with a slight reduction to 23.0% in 2018/2020 (Figure 1).

In bivariate analysis, ESR prevalence was consistently higher among those aged 35-44 than men aged 15-19 years across all the four study periods in 2013/15, 2015/16, 2016/18 and 2018/20 [Table 2].

**Table 2:**
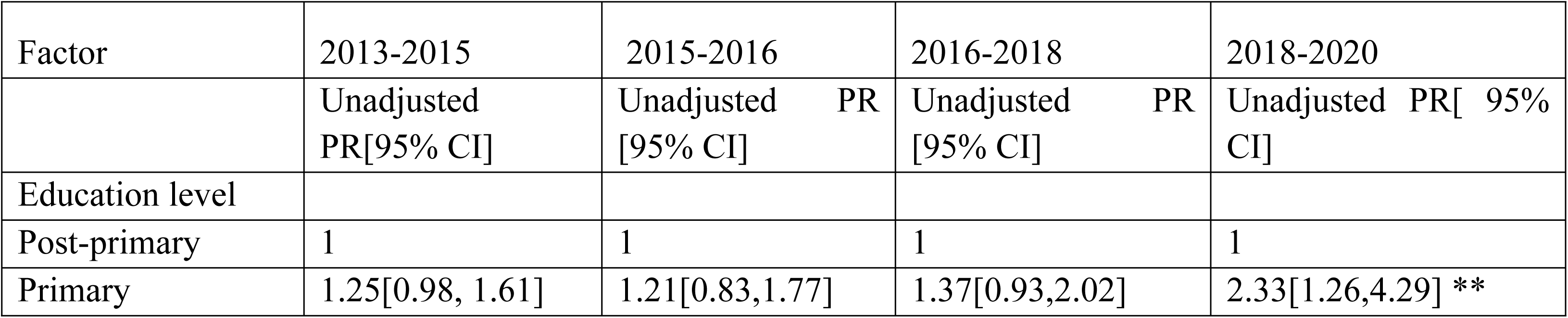
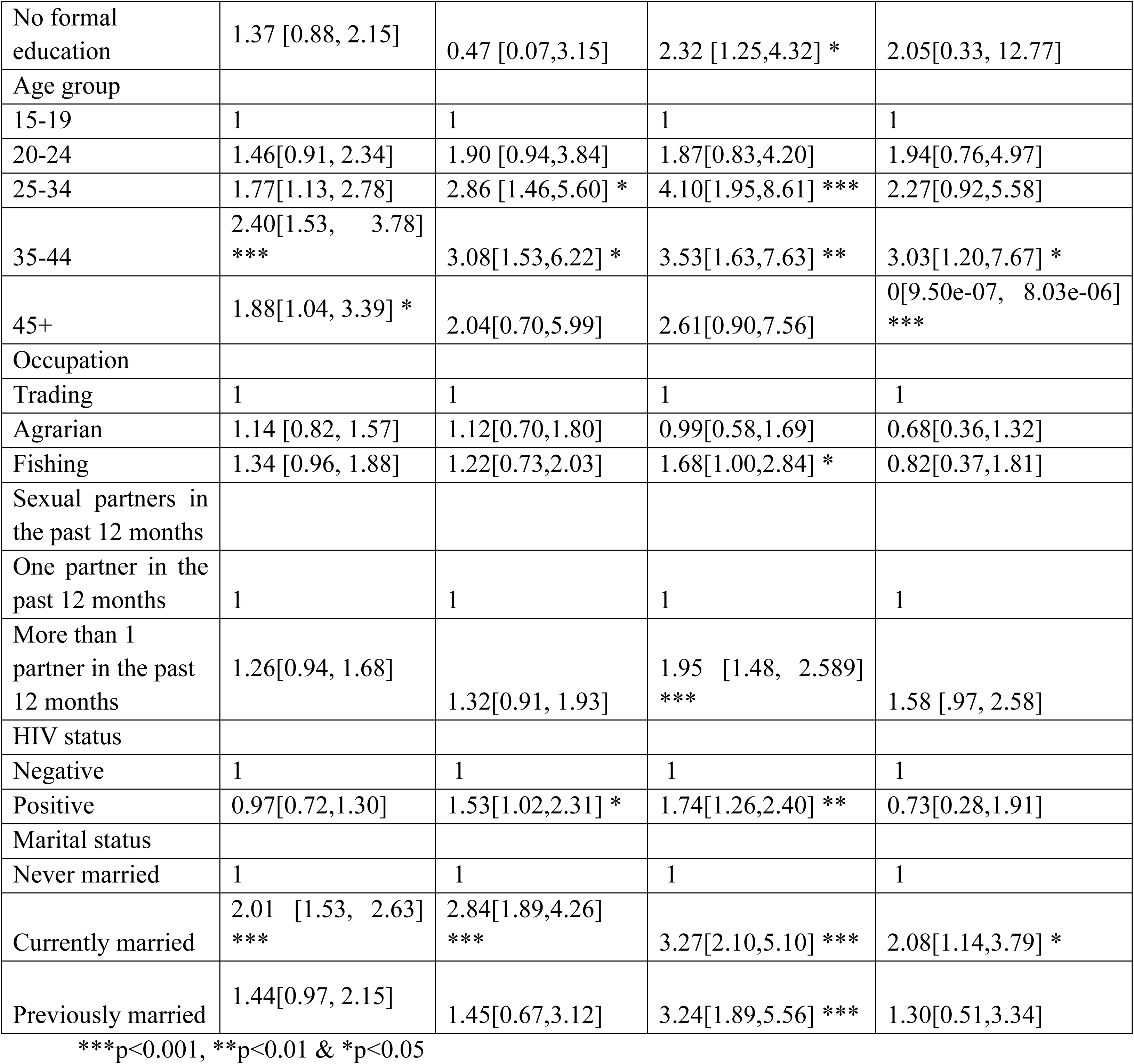
Unadjusted Early Sexual Resumption Prevalence Ratios post-circumcision by survey round 2013-2020 in Rakai, Uganda.

In addition, ESR prevalence was significantly higher among currently married men than men who never married across all surveys (Table 2).

ESR prevalence was significantly higher among HIV-positive men compared to HIV-negative men in 2015/2016 and 2016/2018; PR=1.53, 95% CI: [1.02,2.31]; and PR= 1.74, 95% CI: [1.26,2.40] respectively. However there we no significant difference in rounds 2013/2015 and 2018/2020. ESR was significant in men with multiple sex partners than those reporting one sexual partner in one survey round, 2016/2018 aPR=1.95, 95% CI: [1.48, 2.58]. However, this was not significant in three survey rounds, 2013/15, 2015/16 and 2018/20, aPR=1.26, 95% CI: [0.94, 1.68], aPR=1.32, 95% CI: [0.95, 1.84] and aPR=1.46, 95% CI: [0.83,2.59], respectively. In 2016/2018,

ESR was significantly higher among fisher men compared to traders; PR=1.68, 95% CI: [1.00,2.84] and in 2016/18 ESR was higher among men with no formal education compared to men with post-primary education; PR=2.32 95% CI: [1.25,4.32]. ESR was more prevalent among men with primary education than men with post-primary education; PR=2.33, 95% CI: [1.26,4.29] in 2018/2020.

Multivariate analysis of risk factors associated with ESR across all study periods (2013-2020) Table 3 shows the multivariate analysis, in 2018/2020 and after adjusting for marital status, age, HIV status, occupation, and number of sexual partners in the past 12 months ESR was significantly higher among men who reported primary education as their highest level of completed education than men with post-primary education; aPR=2.38, 95% CI: [1.31, 4.30]. ESR was also significantly higher among men who were currently married than men who never married, and this was consistent across three study periods in 2013/15, 2015/16, 2016/18 and it was not significant in 2018/2020. In 2016/2018, ESR was also significantly higher among unmarried men than men who never married; aPR=1.96, 95% CI: [0.99,3.90]. In 2016/2018, ESR was higher among men who had more than one sexual partner than those who had one sexual partner, aPR=1.76, 95% CI: [1.32,2.34]. However, this finding was not significant across three study survey periods.

**Table 3:** Adjusted Early Sexual Resumption post-circumcision by round 2013-2020 in Rakai, Uganda.

| Factor | 2013/2015 | 2015/2016 | 2016/2018 | 2018/2020 |
| --- | --- | --- | --- | --- |
|  | Adjusted PR (aPR)[95% CI] | Adjusted PR (aPR) [95% CI] | Adjusted PR (aPR) [95% CI] | Adjusted PR (aPR) [95% CI] |
| Education level |  |  |  |  |
| Post-primary | 1 | 1 | 1 | 1 |
| Primary | 1.23[0.97,1.56] | 1.17 [0.80,1.71] | 1.12[0.75,1.68] | 2.38[1.31, 4.30] ** |
| No formal education | 1.25[0.80,1.94] | 0.53[0.08,3.61] | 1.53 [0.85,2.75] | 2.21[0.34, 14.50] |
| Age group |  |  |  |  |
| 15-19 | 1 | 1 | 1 | 1 |
| 20-24 | 1.22 [0.74,2.00] | 1.36[0.65,2.86] | 1.19[0.50, 2.83] | 1.56[0.64, 3.84] |
| 25-34 | 1.16 [0.70,1.94] | 1.37[0.63,2.97] | 1.79[0.70,4.54] | 1.68[0.62, 4.51] |
| 35-44 | 1.55[0.92,2.63] | 1.38[0.60,3.16] | 1.39[0.53,3.64] | 2.17[0.76, 6.15] |
| 45+ | 1.16[0.59,2.25] | 0.95[0.29,3.11] | 0.99[0.29,3.39] | 7.08e-07[1.86e-07 2.69e-06] *** |
| Occupation |  |  |  |  |
| Trading center | 1 | 1 | 1 | 1 |
| Agrarian | 1.05 [0.77,1.44] | 1.25[0.80,1.95] | 1.16[0.69,1.95] | 0.78[0.37, 1.63] |
| Fishing | 1.20[0.86,1.67] | 0.94[0.56,1.58] | 1.30[0.77,2.18] | 0.67[0.28, 1.62] |
| Sexual partners in the past 12 months |  |  |  |  |
| One partner in the past 12 months | 1 | 1 | 1 | 1 |
| More than one partner in the past 12 months | 1.18 [0.88,1.59] | 1.26[0.86, 1.85] | 1.76[1.32,2.34] *** | 1.17 [0.70, 1.96] |
| HIV status |  |  |  |  |
| Negative | 1 | 1 | 1 | 1 |
| Positive | 0.77[0.57,1.05] | 1.24[0.80,1.92] | 1.08 [0.78,1.50] | 0.57 [0.22, 1.47] |
| Marital status |  |  |  |  |
| Never married | 1 | 1 | 1 | 1 |
| Currently married | 1.83[1.30,2.57] ** | 2.46[1.50,4.06] *** | 2.22[1.22,4.03] ** | 1.32[0.61, 2.84] |
| Previously married | 1.29[0.82,2.01] | 1.34[0.59,3.04] | 1.96[0.99,3.90] * | 0.83[0.30, 2.27] |

\*\*\*p<0.001, \*\*p<0.01 & \*p<0.05

ESR was significantly lower among men ≥45 years than men aged 15-19 years in 2018/2020, aPR=0, 95% CI: [9.50e-07, 8.03e-06]. Finally, occupation and HIV status were not associated with ESR across all study survey rounds.

Overall, ESR declined over time (Table 4) from 45.1% in 2013/2015 to 21.8% in 2015/2016; 21.1% in 2016/2018 and 14.9% in 2018/2020.

**Table 4:**
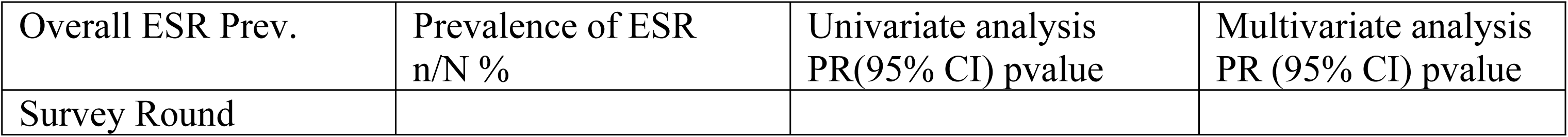
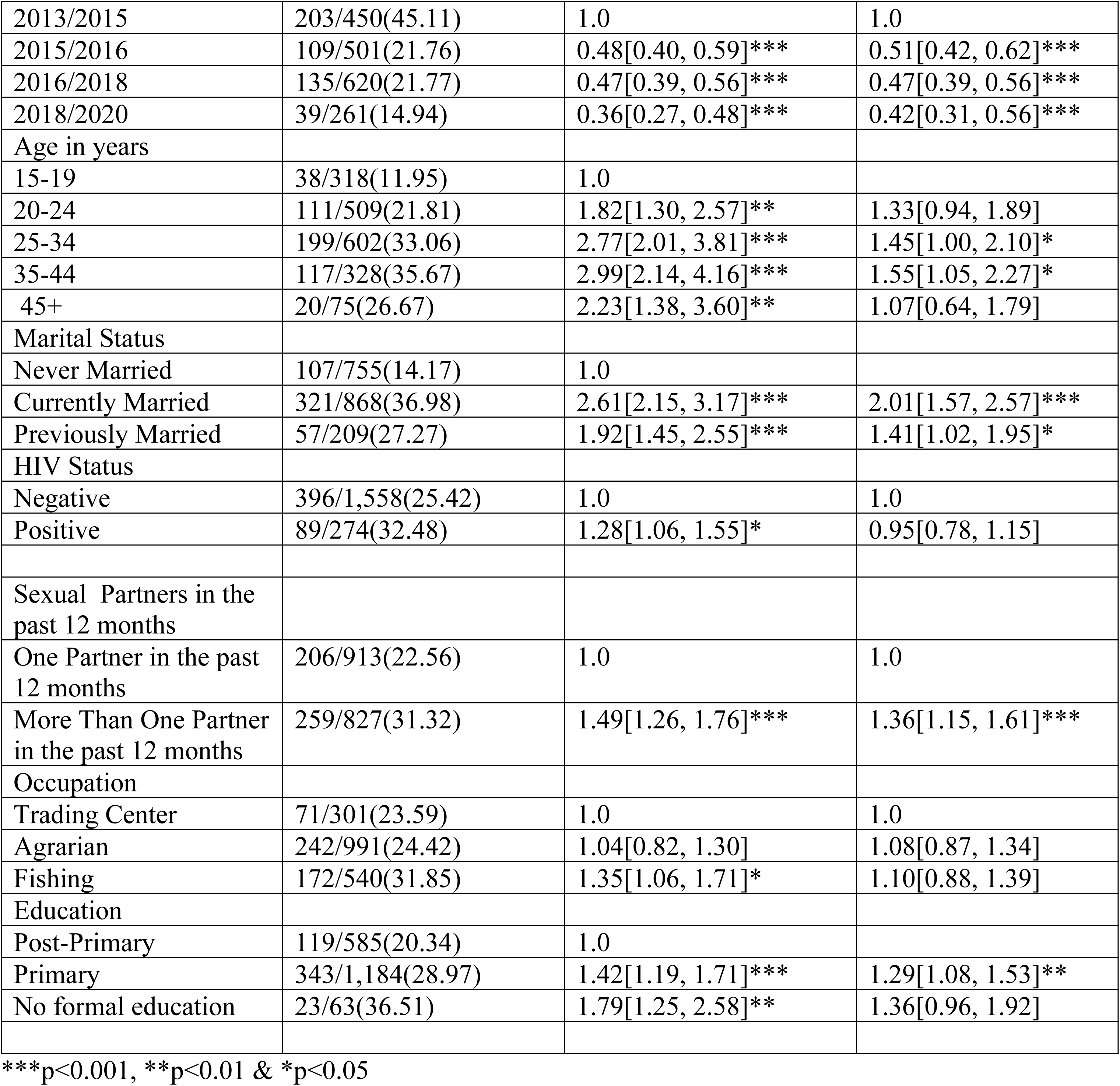
Overall merged ESR Prevalence, Unadjusted and Adjusted Early Sexual Resumption Prevalence Ratios post-circumcision all four surveys (2013-2020) in Rakai, Uganda.

In addition, recent survey visits in 2016/18 and 2018/20, ESR significantly increased, for instance ESR prevalence was significantly higher among men aged 25-44 years than men aged 15-19 years. ESR prevalence was significantly higher among married men and previously married men compared to men who never married. ESR prevalence was significantly higher among men with primary level of education than post-primary aPR=2.38, 95% CI: [1.31, 4.30] in 2018/20. ESR prevalence was significantly higher among men with multiple sexual partners than men without sexual partners aPR=1.36, 95% CI: [1.15, 1.61]

## Discussion

We observed a twofold decline in ESR following VMMC over the 7-year period (2013-2020). A prevalence greater than 35% reported between 2013-2015 is consistent with the prevalence reported in Kenya between 2008 and 2010 [9] and a relative decline in prevalence noted in Uganda between 2015-2016 and 2016-2018 is comparable with an ESR prevalence reported in Kenya [13]. This decline over time could be due to comprehensive HIV programs which emphasized delayed resumption of sex after VMMC that started in 2016 where counseling services were strengthened.[17]. Despite the decline, ESR prevalence remains higher than the recommended WHO target [5]. This puts men at risk of complications and potential HIV acquisition and transmission to female partners [6, 11].

We observed a relationship between marital status and ESR. Married men were consistently more likely to report ESR than unmarried men consistent with at least one other study [8]. This could be partly due to pressure from wives [18]. This result is also consistent with results from a qualitative study where men reported fear of losing their wives as a key driver to them resuming sex early [19]. Therefore, we think that counseling wives about the importance of waiting would be beneficial to prevent early sex resumption.

Additionally, an association was observed between age and ESR following VMMC. Men aged 20-24 years and those aged 25-34 years were more likely to resume sex early compared to those aged 15-19 years. This finding is consistent with previous research conducted in Kenya [8], [9]. The explanation for this association could be because these men are more likely to have multiple sex partners who could be increasing pressure on them to resume sex early. Strong post-circumcision counseling for older men is critical for controlling HIV incidence and prevalence in these populations [20].

Furthermore, participants who had multiple sexual partners were more likely to resume sex early compared to those who had one sexual partner. This finding is consistent with findings from [9], [12]. Health education and sensitization strategies that encourage complete wound healing and other safe sexual practices are recommended for men with multiple sexual partners to benefit from the benefits accrued from VMMC.

A limitation to this study is that sexual resumption questions were based on self-reports and is likely to be under-reported due to social desirability. Secondly, we could not tell if the client was HIV-positive at the time of VMMC service uptake. Further qualitative research to understand client and partner attitudes about early sexual resumption would be of value.

## Conclusions

Generally, ESR among recently-circumcised men declined from 2013 to 2020, suggesting that counseling may have affected post-VMMC behaviors. ESR was more common among married men and men reporting multiple sex partners, suggesting that involvement of female partners could increase compliance with WHO guidelines. Men with lower education also had higher rates of ESR suggesting that targeted counseling for men with lower education could reduce ESR.

## Authors’ contributions

[Mandatory]

AD contributed to conception of this research idea including supporting all stages of this research. FN, EK, AK, BN, GNK, RS,AS, JN, TL, JK, GN, SA, LAM, GK, RG, MW, DS, NS, GK supported with the study conception, design, revisions and editing.

## Data Availability

All data files will be available upon request.

## Acknowledgements

[Mandatory]

We thank Rakai Health Sciences Program (RHSP) for availing the data. We would like to extend our gratitude to our RCCS participants for sparing time and participate in the study.

## Disclaimer

The findings and conclusions in this report are those of the authors and do not necessarily represent the official position of the funding agencies.

